# Intergenerational Persistence of Health: Evidence from India

**DOI:** 10.1101/2023.02.04.23285481

**Authors:** Santosh Kumar, Bernard Nahlen

## Abstract

Using nationally representative data, we estimate intergenerational persistence in health in India. Results from the instrumental variable method show that children of anemic mothers are more likely to be anemic, with an intergenerational health correlation of 0.26. Results are robust to the inclusion of confounding factors including the mother’s height. We find that the correlation between mothers’ anemic status and children’s anemic status differs by wealth quintile, indicating that economic status may play a role in the persistence of poor health across generations in developing countries.

**JEL Classification:** I10, I14, O15

## 1. Introduction

One of the most topical and urgent public health challenges in low- and middle-income countries (LMICs) is micronutrient deficiencies (MND) among children and women. High levels of MND among women and children may signify the existence of intergenerational transmission of poor health from mother to children. Existing work on intergenerational transmission has focused on the labor market (Azam, 2015) and educational outcomes; however, intergenerational transmission of health is understudied and little is known about the independent effect of maternal health on child health after accounting for genetic factors and unobserved omitted factors in a causal framework (Akbulut-Yuksel and Kugler, 2016; Onyeneho et al., 2019; Halliday, Mazumder, and Wong, 2021).

This study examines the intergenerational transmission of anemia in India and explores the role of economic status in moderating the intergenerational cycle of anemia from mother to child. Prior research has examined intergenerational transmission of health in the context of birthweight, height, weight, etc. (Currie and Moretti, 2007; Akbulut-Yuksel and Kugler, 2016), but the evidence on transmission of anemia from mother to child is limited. Identifying the factors that generate intergenerational transmission of health is important because of the high incidence of anemia globally—about 1.6 billion people are affected by anemia, predominantly in Asia and Africa. Two-thirds of LMICs’ anemia cases are in India—an anemia prevalence of 67% among under-five children and 57% among 15-49 years old women (IIPS and ICF, 2021). Furthermore, previous studies related to fetal programming and early-life literature have shown that micronutrient deficiencies in the pre- and post-natal phase of life have adverse effects on health, human capital, labor productivity, and socioeconomic outcomes later in life (Currie and Vogl, 2013; Almond, Currie, and Duque, 2018). A recent modeling study estimates that a scaling-up of antenatal iron-folic acid supplementations could lead to gains of 2.28 million school years and US$8.25 billion in lifetime income (Perumal et al., 2021). Thus, addressing the anemia problem in LMICs would help spur human capital formation and would lead to substantial economic benefits.

Intergenerational health association (IHA) could be due to genetic factors or due to causal effects of parental behaviors on children’s outcomes (Lochner, 2008). Intergenerational transmission of health behaviors and outcomes could be social in nature and is likely to be determined by socioeconomic environments and is not always purely genetic (Lochner, 2008). Identifying the role of genetic, parental, and socioeconomic factors in the IHA is important to understand the effectiveness of policies or factors in breaking the cycle of transmission of poor health from mother to child. Halliday et al. (2021) estimate a health mobility coefficient in the USA with a range of 0.20-0.25. The intergenerational persistence in birth weight ranged from 0.17-0.20 (Currie and Moretti, 2007), and Akbulut-Yuksel and Kugler (2016) show intergenerational association in BMI and height among natives as well as immigrants in the USA. Most of the studies on intergenerational persistence in health are from high-income countries; thus, this study extends intergenerational persistence in micronutrient deficiency in an LMIC setting.

The objective of this study is twofold: to estimate the intergenerational correlation in anemia between mother and child in India, and to explore heterogeneity in this correlation by the household’s economic status after controlling for genetic factors. We accomplish these objectives by using the instrumental variable method to estimate the causal association between the mother’s anemic status and the children’s anemic status. We employ a leave-out-one instrument at the district level and include the mother’s height to further control for the genetic factor. We estimate an intergenerational correlation of 0.40 and 0.26 between mothers’ and children’s Hb levels and anemic status, respectively. This implies that children of anemic mothers have a 26 percentage point higher probability of being anemic. The heterogeneity analyses by wealth quintiles show that economically disadvantaged children display lower upward mobility compared to children from better-off households.

## 2. Data and empirical methodology

We use micro-level data from the fourth round of the National Family Health Survey (NFHS) conducted in 2015-16 (IIPS & ICF, 2017). The NFHS is a nationally representative health survey gathering information on population, health, and nutrition from a sample covering 601,509 households and 699,686 women.

The NFHS survey collected hemoglobin (Hb) information for 6-59 months old children and for 15-49 years old pregnant as well as non-pregnant women. The main dependent variable is the Hb level and anemic status of children born in the last five years before the survey. Children are categorized as anemic if the Hb < 11.0 g/dL while the cutoff for pregnant and non-pregnant women is 11.0 g/dL and 12.0 g/dL, respectively. The anemic category includes severely, moderately, and mildly anemic observations. The control variables include gender, age, and birth order of the child and household-level controls include the mother’s education, mother’s height, religion, social group of the household (caste), and rural dummy. We trim 1 percent of observations at the bottom and top tail of the Hb distribution to exclude outliers. The analytical sample has 198,315 observations.

Table 1 presents the summary statistics for the variables used in the analysis. The average Hb level is 10.59 g/dL and about 58% of the sampled children are anemic. More than half of the women are anemic (56%). The average age of the children is 2 years. Mothers’ mean years of schooling is 6 years and about one-half of the households belong to the bottom two wealth groups. More than two-thirds of the children reside in rural areas (76%).

**Table 1:** Descriptive statistics for analytical sample (N= 198,203)

| **Variables** | **Mean** | **St. Dev.** |
| --- | --- | --- |
| <u>Outcomes</u> |  |  |
| Child Hb level | 10.59 | 1.38 |
| Mother Hb level | 11.52 | 1.47 |
| Child is anemic (%) | 0.58 | 0.50 |
| Mother is anemic (%) | 0.56 | 0.50 |
| District anemia rate (%) | 0.56 | 0.12 |
| <u>Child demographics</u> |  |  |
| Female (%) | 0.48 | 0.50 |
| Birth order | 2.26 | 1.40 |
| Age | 2.23 | 1.30 |
| <u>Households' demographics</u> |  |  |
| Mother's education | 6.07 | 5.11 |
| Religion | 0.73 | 0.45 |
| Access to improved toilet | 0.51 | 0.50 |
| Social group (SC-ST) | 0.39 | 0.49 |
| <u>Wealth groups</u> |  |  |
| Poorest | 0.27 | 0.44 |
| Poor | 0.24 | 0.24 |
| Middle | 0.20 | 0.39 |
| Rich | 0.16 | 0.37 |
| Richest | 0.13 | 0.34 |
| Rural (%) | 0.76 | 0.42 |
| Number of districts | 628 |  |
| Number of states | 35 |  |

Notes: SC-ST: Scheduled caste and scheduled tribe; mother's education is in years of schooling

Following the standard method in this literature (Halliday et al., 2020), we estimate the following linear regression model to estimate the intergenerational health association between mother and children:

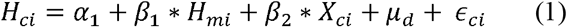

Where *H*_*ci*_ denotes the health of the child in the family i; *H*_*mi*_ denotes the health of the mother *m*; *X*_*Ci*_ denotes a vector of mothers’ and children’s level controls; *µ*_*d*_ is district fixed effects to control for time-invariant characteristics of the district. Standard errors are clustered by the district. We weight the regression models with national women’s sample weight.

Since the main independent variable, the mother’s anemic status, is likely to be endogenous, we employ the Two-Stage Least Square (2SLS) regression model to address endogeneity. For example, unobserved heterogeneity such as the mother’s health, dietary behavior, and genetic factors may affect the mothers’ as well as children’s anemic status, and therefore, the OLS estimates would be biased and inconsistent. Furthermore, mothers and their children share a similar socioeconomic environment and that may affect mothers’ and children’s health. To address this endogeneity problem, we instrument mothers’ anemic status by the leave-out average anemia rate among rural women at the district level. A similar instrument i.e. endogenous variable aggregated at the district or state level has been used in prior studies (Fruehwirth et al., 2019).

The first-stage regression equation is:

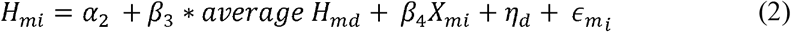

And the predicted value of the mother’s anemia from equation (2) is used in the second stage:

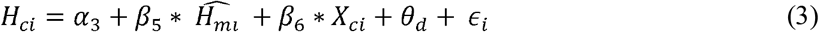

The parameter *β*_5_ is the measure of intergenerational persistence of health between mothers and children. 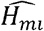 is the predicted value of mother’s health from equation (2). The instrument relevance condition is satisfied when the district-level average anemia rate is strongly correlated with the mother’s anemic status. The exclusion restriction is not testable, but it is reasonable to argue that the exclusion restriction is likely to be satisfied after including a robust set of controls and district-fixed effects.

## 3. Findings

Table 2 shows that after adjusting for the mother’s height, the mother’s anemic status is statistically significantly associated with the anemic status of their children. If a mother is anemic, the anemia probability for children increases by 10.4 percentage points (pp) (col 4). The intergenerational correlation in Hb is 0.14.^2^

**Table 2:** Intergenerational correlation in Hb and anemia, OLS results

|  | Child's Hb level |  | Child is anemic |  |
| --- | --- | --- | --- | --- |
|  | (1) | (2) | (3) | (4) |
| Mother's Hb level | 0.138***<br>(0.003) | 0.143***<br>(0.003) |  |  |
| Mother is anemic |  |  | 0.104***<br>(0.004) | 0.104***<br>(0.004) |
| Mother's height | No | 0.003***<br>(0.0008) | No | -0.00009***<br>(0.00003) |
| Child controls | Yes | Yes | Yes | Yes |
| Household controls | Yes | Yes | Yes | Yes |
| District fixed effects | Yes | Yes | Yes | Yes |
| Observations | 198,203 | 198,203 | 198,203 | 198,203 |

Notes: Hb- hemoglobin level in grams per deciliter (gm/dL). Robust standard errors, clustered at the district level, are reported in parentheses. Child controls include gender, birth order, and age, while household controls include the mother's education, religion, caste, wealth index, and rural dummy.
\* Indicates significance at 10% level.
\*\* Significance at 5% level.
\*\*\* Significant at 1% level of confidence.

Our preferred 2SLS estimates are presented in Table 3. The first-stage results show that the instrument is statistically significantly correlated with the anemic status of the mother. The *F*-statistics on the excluded instrument is more than 10 (Stock and Yogo, 2002), indicating that the instrument is strongly correlated with the endogenous variable and that the instrument does not suffer from weak instrument problems. Results further show that the instrument passes the weak-identification test and is robust to weak instrument inference. The 2SLS estimates are positive and statistically significant. Col (2) shows that when a mother’s Hb level increases by one unit (1 g/dL), children’s Hb level increases by 0.40 g/dL. Similarly, children of the anemic mother are 26 pp more likely to be anemic (col 4). The 2SLS estimates are almost 2.5 times larger than the OLS estimates, suggesting that the OLS estimates underestimate the true intergenerational correlation between mothers’ and children’s health. The OLS and 2SLS estimates cannot be compared directly because the two estimates refer to different population groups. The 2SLS estimate gives the local average treatment effect (LATE) for a subgroup of the population, while OLS estimates the average treatment effect (ATE) for the whole population. LATE can be greater than ATE if there are heterogeneous treatment effects. For the results presented here, the IHA correlation is larger for the LATE subgroup than the whole population.

**Table 3:** Intergenerational correlation in Hb and anemia, 2SLS results

|  | Child's Hb level |  | Child is anemic |  |
| --- | --- | --- | --- | --- |
|  | (1) | (2) | (3) | (4) |
| *First-stage results* |  |  |  |  |
| District anemia rate | -2.43***<br>(0.131) | -2.43***<br>(0.131) | 0.967***<br>(0.020) | 0.968***<br>(0.020) |
| F-test | 344.56 | 345.89 | 2289.0 | 5.67 |
| *Weak identification test* |  |  |  |  |
| Cragg-Donald Wald F statistic | 767.58 | 769.90 | 1221.41 | 1222.81 |
| Kleibergen-Paap Wald rk F statistic | 344.56 | 345.89 | 2289.0 | 2282.39 |
| *Weak-instrument-robust inference* |  |  |  |  |
| Anderson-Rubin Wald test (p-value) | <0.001 | <0.001 | <0.001 | <0.001 |
| Stock-Wright LM S statistic | <0.001 | <0.001 | <0.001 | <0.001 |
| *Two-stage least square (2SLS) results* |  |  |  |  |
| Mother's Hb level | 0.397***<br>(0.065) | 0.395***<br>(0.065) |  |  |
| Mother is anemic |  |  | 0.263***<br>(0.046) | 0.262***<br>(0.046) |
| Mother's height | No | 0.0003***<br>(0.00008) | No | -0.00009***<br>(0.00003) |
| Child controls | Yes | Yes | Yes | Yes |
| Household controls | Yes | Yes | Yes | Yes |
| District fixed effects | Yes | Yes | Yes | Yes |
| Observations | 198,203 | 198,203 | 198,203 | 198,203 |

Notes: Hb- hemoglobin level in grams per deciliter (gm/dL). Robust standard errors, clustered at the district level, are reported in parentheses. Child controls include gender, birth order, and age, while household controls include the mother's education, religion, caste, wealth index, and rural dummy. \* p < 0.10, \*\* p < 0.05, \*\*\* p < 0.01

Previous studies have shown that maternal height captures the genetic factors and affects the nutrition-related intergenerational transmission of health, therefore, it is important to control for maternal height in the empirical model. The coefficients for the mother’s height are positive and statistically significant, but the effect size is small. The results in Table 2 show that the inclusion of the mother’s height does not change the IHA coefficient much, indicating a muted role of genetic factors in the transmission of health across generations. Our results allude to a greater role of socio-economic environments in shaping the correlation between mother and child anemia since mothers and children share the same socio-economic environment.

Furthermore, the subgroup analysis by wealth index shows that the IHA correlation in Hb levels and any form of anemia is substantially larger for the bottom two wealth quintiles compared with the top two wealth quintiles (Fig. 1). The detailed regression results are reported in Table A1. The pattern of wealth gradient in Fig. 1 indicates that children are more likely to be anemic if they are born in households with lower economic resources. This suggests that improvement in the economic status of the household will limit the extent to which child health is tied to mother health. Government programs designed to improve the economic conditions of populations may break the intergenerational transmission of poor health across generations. Given the confidence interval overlap, our findings related to the wealth quintile, while interesting, should be interpreted with caution.^3^

**Fig. 1:**
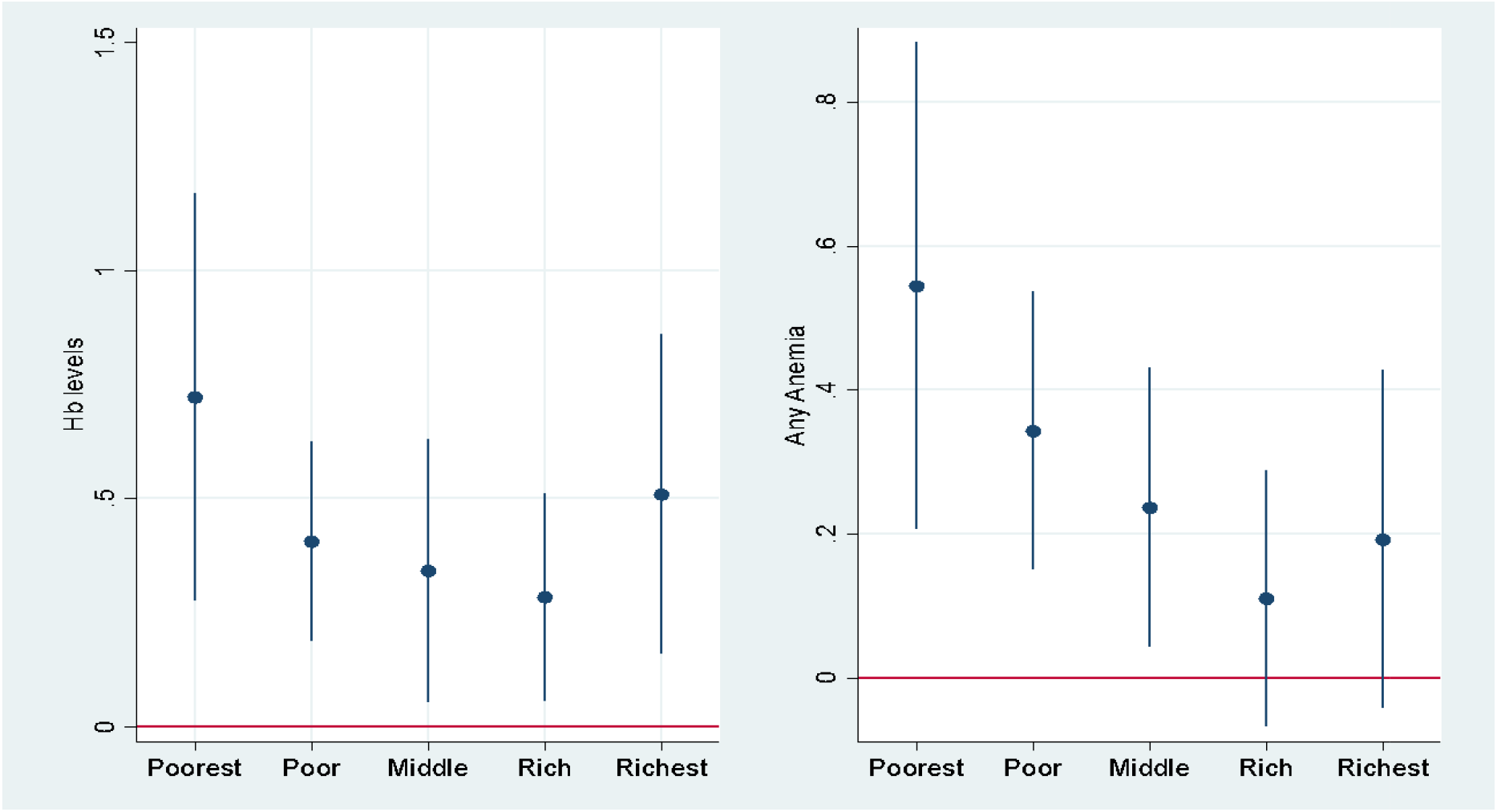
Heterogeneous effects by wealth index, 2SLS estimates Notes: Child controls include gender, birth order, and age, while household controls include the mother’s education, mother’s height, religion, caste, mother’s height, and rural dummy. Using principal component analysis, the wealth index is calculated based on a household’s ownership of selected assets, such as televisions and bicycles; materials used for housing construction; and types of water access and sanitation facilities. Households are divided into quintiles from the poorest to the richest.

## 4. Conclusion

This paper is a handful of studies to investigate the IHA between mother and child health in a causal framework in India, except the study by Onyeneho et al., (2019). Analyzing large and nationally representative data, we find a high degree of intergenerational persistence in anemia. The study contributes to limited evidence on IHA in developing countries and in the context of anemia which is not self-reported and, therefore, is less likely to suffer from measurement errors. There could be some concerns about the exclusion restriction, for example, micronutrients in the crops grown in the region may affect the instrument as well as the outcome variable; therefore, our 2SLS estimates should be interpreted with caution. These are important findings that highlight the need to address anemia and other health issues in a comprehensive and targeted manner, particularly among disadvantaged populations from lower wealth groups. Future studies should consider other potential factors that may contribute to the persistence of anemia and other health problems across generations, including access to quality health care, nutrition, religion, gender, and other social and economic determinants of health.

## Data Availability

All data produced are available online at https://dhsprogram.com

## Data availability

The authors do not have permission to share data.

## Acknowledgment

The authors thank the editor, Prof. Audra Bowlus, an anonymous referee, and Timothy Halliday for valuable suggestions.

## Appendix

### See Table A1

**Table A1:**
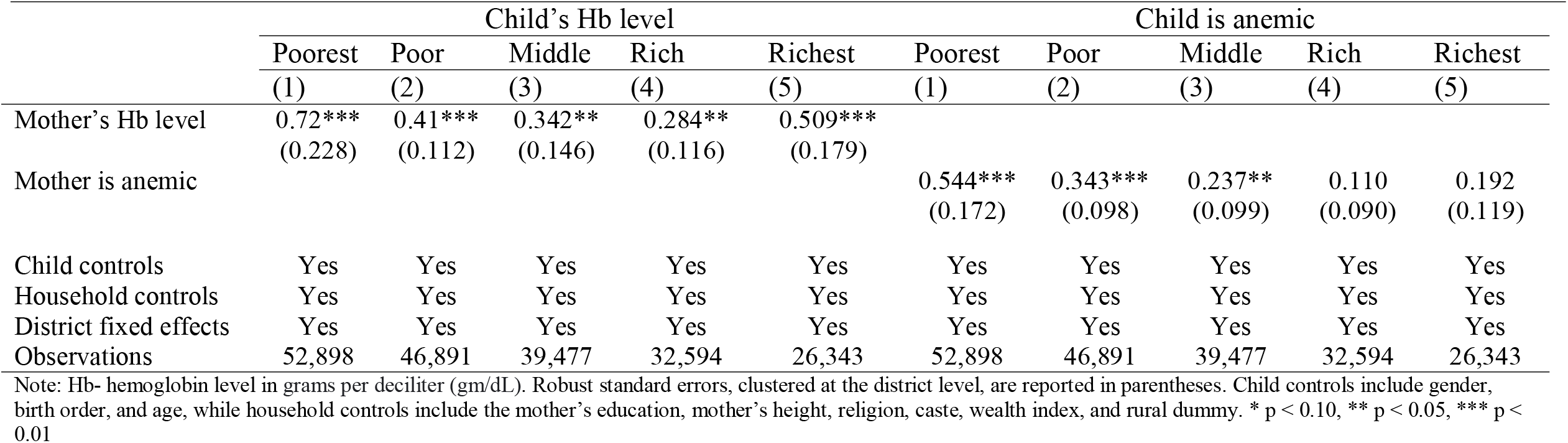
Heterogeneous effects by wealth index, 2SLS estimates

|  | Child's Hb level |  |  |  |  | Child is anemic |  |  |  |  |
| --- | --- | --- | --- | --- | --- | --- | --- | --- | --- | --- |
|  | Poorest | Poor | Middle | Rich | Richest | Poorest | Poor | Middle | Rich | Richest |
|  | (1) | (2) | (3) | (4) | (5) | (1) | (2) | (3) | (4) | (5) |
| Mother's Hb level | 0.72***<br>(0.228) | 0.41***<br>(0.112) | 0.342**<br>(0.146) | 0.284**<br>(0.116) | 0.509***<br>(0.179) |  |  |  |  |  |
| Mother is anemic |  |  |  |  |  | 0.544***<br>(0.172) | 0.343***<br>(0.098) | 0.237**<br>(0.099) | 0.110<br>(0.090) | 0.192<br>(0.119) |
| Child controls | Yes | Yes | Yes | Yes | Yes | Yes | Yes | Yes | Yes | Yes |
| Household controls | Yes | Yes | Yes | Yes | Yes | Yes | Yes | Yes | Yes | Yes |
| District fixed effects | Yes | Yes | Yes | Yes | Yes | Yes | Yes | Yes | Yes | Yes |
| Observations | 52,898 | 46,891 | 39,477 | 32,594 | 26,343 | 52,898 | 46,891 | 39,477 | 32,594 | 26,343 |

Note: Hb- hemoglobin level in grams per deciliter (gm/dL). Robust standard errors, clustered at the district level, are reported in parentheses. Child controls include gender, birth order, and age, while household controls include the mother's education, mother's height, religion, caste, wealth index, and rural dummy. \* $p < 0.10$ , \*\* $p < 0.05$ , \*\*\* $p < 0.01$

## Footnotes

2 Similar findings were reported in Onyeneho et al., (2019). The authors find a Hb correlation of 0.1 and 0.2 for father-child and mother-child samples, respectively.

3 It should be noted that overlapping confidence intervals is not an appropriate test of equality because this method does not account for correlation between the estimators, nor does it account for the multiple tests performed. (Schenker and Gentleman, 2001). It is quite possible that coefficients in Fig. 1 are statistically different despite the overlap in the confidence intervals.

## Notes

### Competing Interest Statement

The authors have declared no competing interest.

### Funding Statement

This study did not receive any funding.

